# Computed tomography perfusion parameters predictive of symptomatic intracranial hemorrhage after mechanical thrombectomy in patients with cerebral large vessel occlusion

**DOI:** 10.1101/2023.02.16.23286072

**Authors:** Soichiro Abe, Manabu Inoue, Manabu Shirakawa, Kazutaka Uchida, Kiyofumi Yamada, Yoji Kuramoto, Satoshi Namitome, Seigo Shindo, Fumihiro Sakakibara, Junichi Kouno, Kotaro Tatebayashi, Norito Kinjo, Shoichiro Tsuji, Shuichi Tanada, Mikiya Beppu, Hidetoshi Matsukawa, Masafumi Ihara, Shinichi Yoshimura

## Abstract

**Background:** Hemorrhagic transformation (HT) after recanalization is a severe complication in patients with a large ischemic core due to cerebral large vessel occlusion. Risk assessment using perfusion imaging to predict hemorrhagic infarction has not been established. Thus, we aimed to investigate the association between perfusion imaging findings and HT in patients with acute cerebral large vessel occlusion who had undergone preoperative perfusion imaging evaluation and mechanical thrombectomy.

**Methods:** We enrolled consecutive patients who received mechanical thrombectomy (MT) after undergoing perfusion imaging for anterior large vessel occlusion from May 2019 to March 2022. The patients in whom recanalization was not achieved and who experienced procedure-related bleeding were excluded. We investigated the predictors of symptomatic intracranial hemorrhage (sICH) by exploring preoperative perfusion imaging parameters [relative cerebral blood flow, relative cerebral blood volume (rCBV), time of maximum concentration, hypoperfusion index ratio].

**Results:** Among the 167 patients (median age 79 years, 47% female) enrolled, 63 (38%) and 14 (8%) patients had any intracranial hemorrhage and sICH (sICH group), respectively. The sICH group had a shorter puncture-recanalization time than the non-sICH group (median [interquartile range (IQR)]; 43 [34–55] vs. 61 [37–88], p = 0.046), whereas the modified Rankin Scale at 90 days showed a worse prognosis (median [IQR]; 5 [5–6] vs 3 [1–4], p<0.01). All perfusion imaging parameters were significantly predicting the sICH group in multiple logistic regression analysis. The value of rCBV was the parameter most strongly associated with sICH in receiver operating characteristic curve analysis (area under the curve = 0.90, 95% confidence interval [0.83–0.98]; cutoff 43 ml; sensitivity, 86 %; specificity, 87%).

**Conclusion:** Among perfusion CT parameters, rCBV is highly associated with sICH after MT for cerebral large vessel occlusion. In patients with low rCBV regions, the indication of mechanical thrombectomy should be carefully considered for postoperative intracranial hemorrhage.

**Clinical perspective:**

- Of the various parameters assessed via perfusion CT, the value of rCBV demonstrates a strong correlation with the occurrence of symptomatic intracranial hemorrhage following mechanical thrombectomy for cerebral anterior large vessel occlusion.
- When treating patients with regions of large low-rCBV, it is important to carefully determine the indication of mechanical due to the potential risk of postoperative intracranial hemorrhage.

## Introduction

Mechanical thrombectomy (MT) for acute cerebral large vessel artery occlusion has been established in the last decade, and has some of the highest levels of evidence among therapies in most countries’ guidelines^1,2^. Two randomized controlled trials (RCTs) were conducted for patients with an unknown time of stroke onset, and it is recommended that the indication of MT be expanded according to tissue-based decision^1,3,4^. Moreover, RESCUE-Japan LIMIT study, which is the RCT on the efficacy of MT for low Alberta Stroke Program Early CT Score, reported that functional outcome was improved in the group treated with MT, even in patients with large ischemic changes^5^. On the other hand, in the patients with a large ischemic core, the risk of hemorrhagic transformation (HT) is high after reperfusion therapy, which might worsen the prognosis^6-8^. Although the incidence of HT is reported to vary widely, ranging from 2% to 44%^9,10,11^, in patients receiving reperfusion therapy including thrombolytic therapy and endovascular therapy, HT is found in two-fifths of these patients after MT^12,13^. Therefore, clarifying the factors causing HT after reperfusion therapy is urgently needed. Factors that are previously reported to predict HT are thrombolytic therapy, younger age^14^, hyperglycemia^15^, low levels of coated platelet^16^, cardiac subtype^17^, acute hypertension^18^, and blood pressure variability^19^. However, the prediction of HT using perfusion imaging has not been established, and it is unknown which parameters can be utilized. Thus, we decided to retrospectively investigate the association between perfusion imaging findings and HT on patients with acute cerebral large vessel occlusion who underwent preoperative perfusion imaging evaluation and MT.

## Methods

The data that support the findings of this study are available from the corresponding author upon reasonable request and after permission from the ethics committees. The reporting of this study adheres to the standards outlined in the Strengthening the Reporting of Observational Studies in Epidemiology (STROBE) statement.

This single-center retrospective cohort study analyzed consecutive patients with large vessel occlusion who presented to our hospital between May 2019 and March 2022. We analyzed the patient’s characteristics at admission, non-contrasted and contrasted CT, including perfusion imaging before reperfusion therapy, and intraoperative angiographic findings. We graded the ischemic core in the middle cerebral artery territory using Alberta Stroke Program Early Computed Tomographic Score (ASPECTS) on CT^20^. Neurological findings were evaluated using National Institutes of Health Stroke Scale (NIHSS) at admission and discharge. The functional outcome was evaluated pre-stroke using the modified Rankin Scale (mRS) and at 3 months after stroke onset. Non-contrasted CT was performed at 36 hours (±12 hours) after MT to evaluate the hemorrhagic changes. According to the European Cooperative Acute Stroke Study II (ECASS II), we identified the hemorrhagic changes as hemorrhagic infarction (HI) 1, HI 2, parenchymal hematoma (PH) 1, and PH 2^21^. The following patients were excluded from the analysis: 1) those who did not undergo CT perfusion; 2) with non-adequate perfusion imaging; 3) recanalization was not achieved after MT [modified Thrombolysis in Cerebral Infarction (mTICI) = 0, 1]; and 4) cerebral hemorrhage occurred due to an intraoperative procedure.

RAPID software (iSchemaView, Menlo Park, CA) was used to analyze perfusion imaging. We measured the following parameters: time of maximum concentration (Tmax) > 4 sec, Tmax > 6 sec, Tmax > 8 sec, and Tmax > 10 sec; relative cerebral blood flow (rCBF) < 20%, rCBF < 30%, rCBF < 34%, and rCBF < 38%, relative cerebral blood volume (rCBV) < 34%, rCBV < 38%, and rCBV < 42%^22^. The regions with rCBF <30% were defined as an ischemic core. The region of difference in rCBF < 30 % and Tmax > 6 sec was defined as an ischemic penumbral area^23^. The ratio of Tmax > 6 sec to Tmax > 10 sec (Tmax6/Tmax10) was defined as the hypoperfusion intensity ratio (HIR)^24,25^.

After CT perfusion, intravenous thrombolysis (at 0.6 mg/kg dose according to Japanese guidelines^26^) was administered in the indicated patients. MT was performed using aspiration catheters and/or stent retrievers approved in Japan during the study period. If stenotic lesions remained, intracranial percutaneous transluminal angioplasty was performed as appropriate, and local intraarterial fibrinolysis was added for the distal lesions. All of these procedures were performed under the board certified neurointerventionist’s decision. The mTICI scale was used as a measure of recanalization.^27^ Additionally, each time course was recorded for stroke onset, visit time (door), puncture, and reperfusion time.

Non-contrasted head CT was performed immediately after surgery and at 36 hours (±12 hours) after surgery to evaluate any hemorrhagic changes. For postoperative neurological findings and functional outcomes, the patients were evaluated for NIHSS and mRS at discharge. After discharge, mRS was evaluated at 90 days after stroke onset (mRS at 90 days) at the outpatient clinic. Patients who were unable to visit at the outpatient clinic were contacted via telephone-interview, and mRS at 90 days was evaluated.

We retrospectively assessed the incidence of symptomatic intracranial hemorrhage (sICH) at 36 hours after MT. sICH was defined as postoperative intracranial hemorrhage with worsening of NIHSS score 4 or higher^28^. The incidence of asymptomatic intracranial hemorrhage (a-sICH), any intracranial hemorrhage (any ICH), including sICH and a-sICH at 36 (±12 hours) after MT was also investigated, and we compared postoperative neurological and functional outcomes. At discharge, we diagnosed the etiology of infarction according to the Trial of Org 10172 in Acute Stroke Treatment (TOAST) criteria^29^.

Patient background factors, and imaging and neurological findings were all retrospectively extracted from medical record information based on the Hyogo Medical University Study (ethical number 202105-103). The institutional review boards approved the opt-out method in place of written informed consent. Neurological evaluation and functional prognosis were obtained by the physician in charge from the patients or their families during hospitalization or at the outpatient clinics. Two board certified neurointerventionists (S.A. and M.S.) evaluated the patients for sICH. The hemorrahgic change was blindly assessed without perfusion imaging. The data supporting this study’s findings can be accessed from the corresponding author upon reasonable request.

Statistical software used for the analysis was R ver. 4.1.3. Continuous variables are presented as the median and interquartile range (IQR), and categorical variables are presented as numbers and percentages. Continuous variables are tested for the differences in bivariates using Student’s t-test or the Wilcoxon rank-sum test and nominal variables were tested for differences in bivariates using Fisher’s exact analysis. Categorical variables are examined using the chi-square test. Patient background factors, neurological evaluation, and functional prognosis were compared between the non-sICH and sICH groups, and between the no-ICH, a-sICH, and s-ICH groups. The missing values are omitted from the analysis. Univariate and multivariate analyses (adjusted by age, intravenous thrombolytic therapy^14^, number of passes, and ASPECTS) were performed on cases with sICH and each perfusion imaging parameter to examine the association between CT perfusion parameter and sICH. Furthermore, receiver operating characteristics (ROC) curve analysis was performed to measure the sensitivity and specificity of each parameter of CT perfusion to sICH. The ROC analysis was used to obtain the Youden Index cutoff point of perfusion volume to discriminate sICH^30^. For each ROC analysis, we compared the ROCs using the Bootstrap method to identify the parameters more closely related to sICH^31^. We compared the sICH with clinical information in the group of patients who were assessed to be at low risk based on CT perfusion parameters.

## Results

Among the 261 patients who underwent MT during the study period, we excluded 23 patients who had posterior circulation artery occlusion (basilar artery occlusion, 15 patients; posterior cerebral artery occlusion, 6 patients; vertebral artery occlusion 2 patients), 31 patients in whom perfusion imaging could not be performed, 24 patients in whom perfusion imaging was inaccurately imaged due to the motion artifact, venous injection failure, and severe heart failure, 11 patients in whom recanalization was not achieved (mTICI 0, and 1), and five patients in whom hemorrhagic changes occurred due to the intraoperative procedure (Figure 1). The final analysis included 167 patients (78 women [47.0%], median [IQR] age of 79 [71–83] years). The patient background characteristics are shown in Table 1. The median pre-stroke mRS was 0 [0–3], and 69 (41%) patients had received oral antiplatelet or anticoagulant agent before admission. Altogether, 103 patients (62%) had hypertension, 68 patients (41%) had hyperlipidemia, and 31 patients (19%) had diabetes mellitus. The number of patients with atrial fibrillation was 85 (51%). The perioperative time course was as follows: 181 [101–288] minutes for onset to door, 53 [40–81] minutes for door to puncture, and 59 [36–83] minutes for puncture to reperfusion. The TOAST classifications at discharge were large-artery atherosclerosis (LAA) in 40 (24%) patients, cardiac embolism (CE) in 93 (56%) patients, other determined embolisms in 32 (19%) patients, and undetermined causes in 2 (1%). Of the 167 patients, any ICH after procedure was observed in 63 patients (38%), and 14 (8%) had sICH. The puncture to reperfusion time was significantly shorter in the sICH group than the non-sICH group (median [IQR]; 43 [34–55] vs. 61 [37–88], p = 0.046), nonetheless, the mRS at 3 months was significantly higher in the sICH group (median [IQR]; 5 [5–6] vs 3 [1–4], p < 0.01).

**Table 1.**
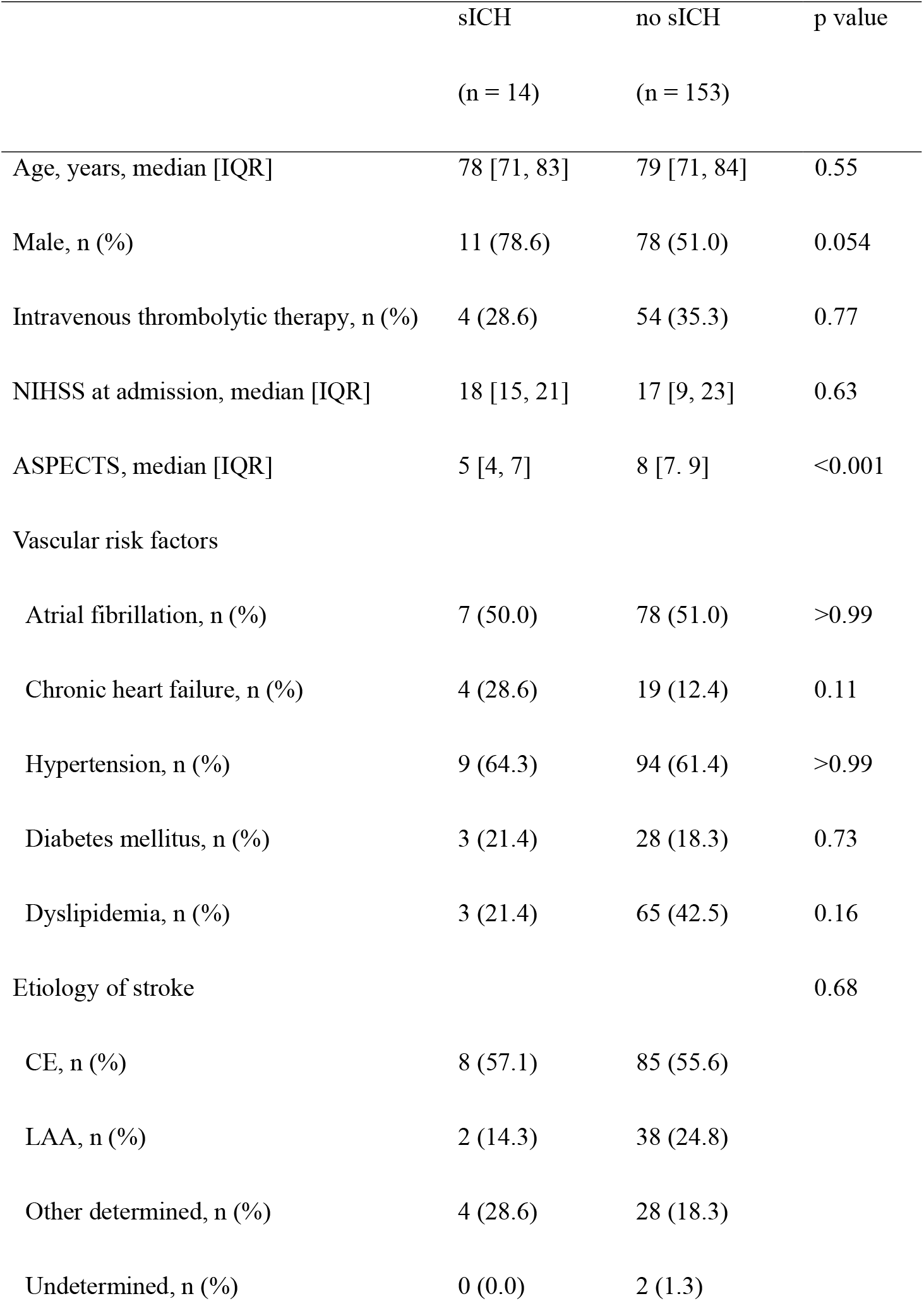
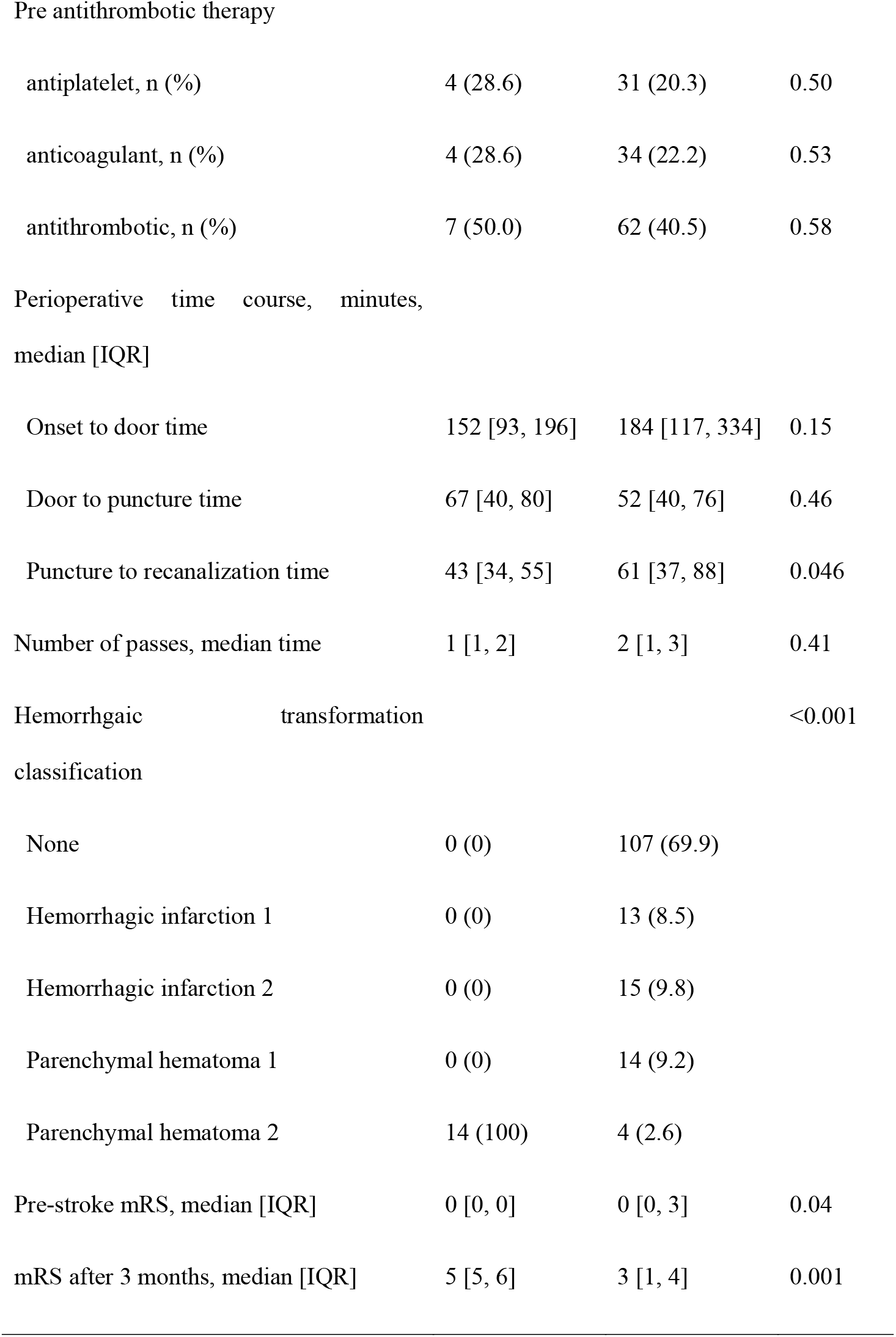
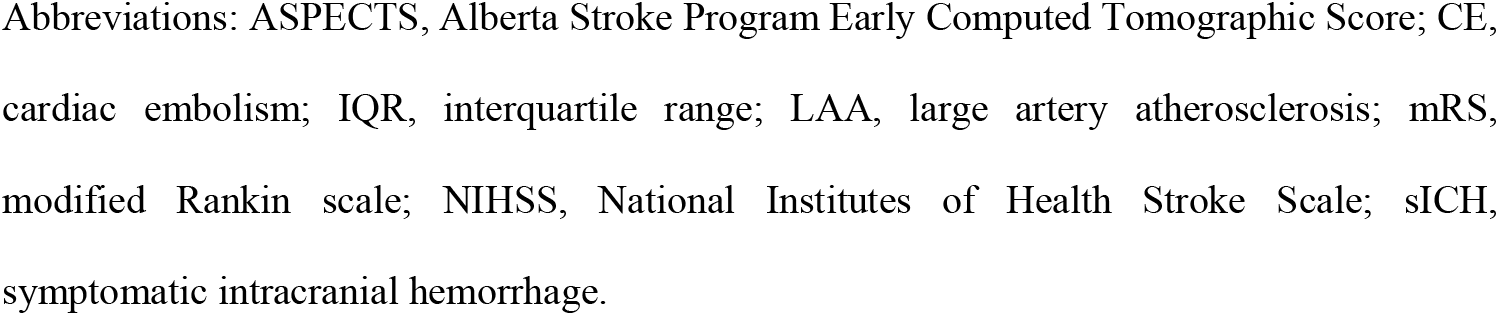
Patient characteristics at baseline; patients with symptomatic intracranial hemorrhage and no symptomatic intracranial hemorrhage.

**Figure 1.**
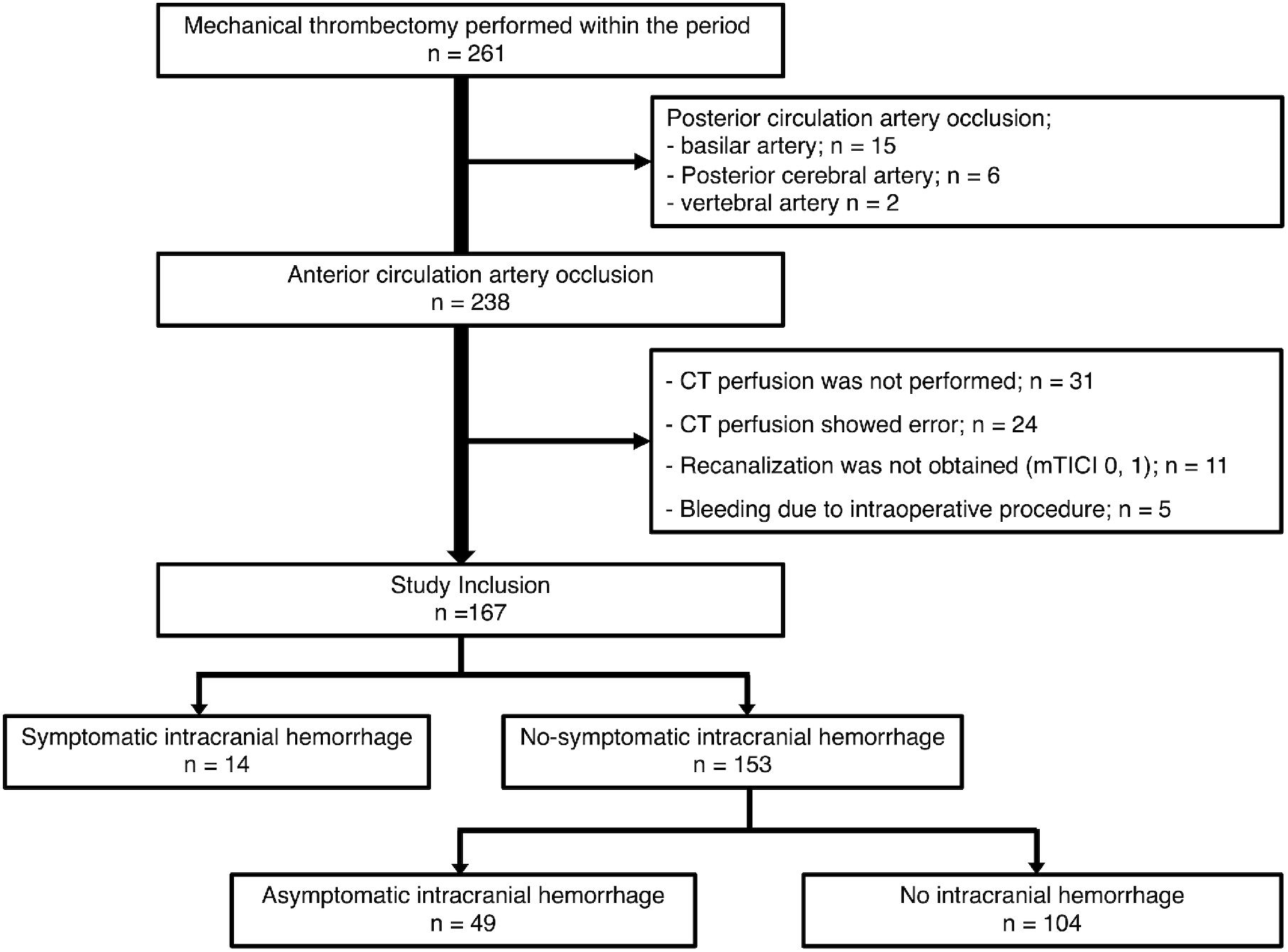
Study flow chart. Flowchart of patient participation in the study. Abbreviations: mTICI, modified Thrombolysis in Cerebral Infarction.

When comparing no-ICH, a-sICH, and sICH groups, the puncture-to-reperfusion time was longer and pre-stroke mRS was higher in the a-sICH group than in the other two groups, and mRS at 3 months was significantly higher in the sICH and a-sICH group than in no-ICH group (Supplemental Table 1).

The results of perfusion imaging evaluated by automated software are shown in Table 2. The parameter of Tmax, rCBF, rCBV, and HIR were all significantly associated with the presence of sICH after adjusting the models. In the ROC analysis, the area under the curve (AUC) of Tmax > 4 sec, Tmax > 6 sec, Tmax > 8 sec, Tmax > 10 sec, rCBF < 30%, rCBV < 34%, HIR were 0.70, 0.77, 0.80, 0.82, 0.87, 0.90, and 0.79, respectively (Figure 2, Supplemental Table 2). To determine the threshold, we compared each parameter and found that the AUC was significantly higher in rCBV < 34% than in rCBF < 30%, Tmax > 10 sec, and HIR (p = 0.01, 0.041, and 0.01, respectively). The rCBV < 34% predicts sICH with a cutoff value of 43 ml with a sensitivity of 86% and specificity of 87% (Figure 2).

**Table 2.**
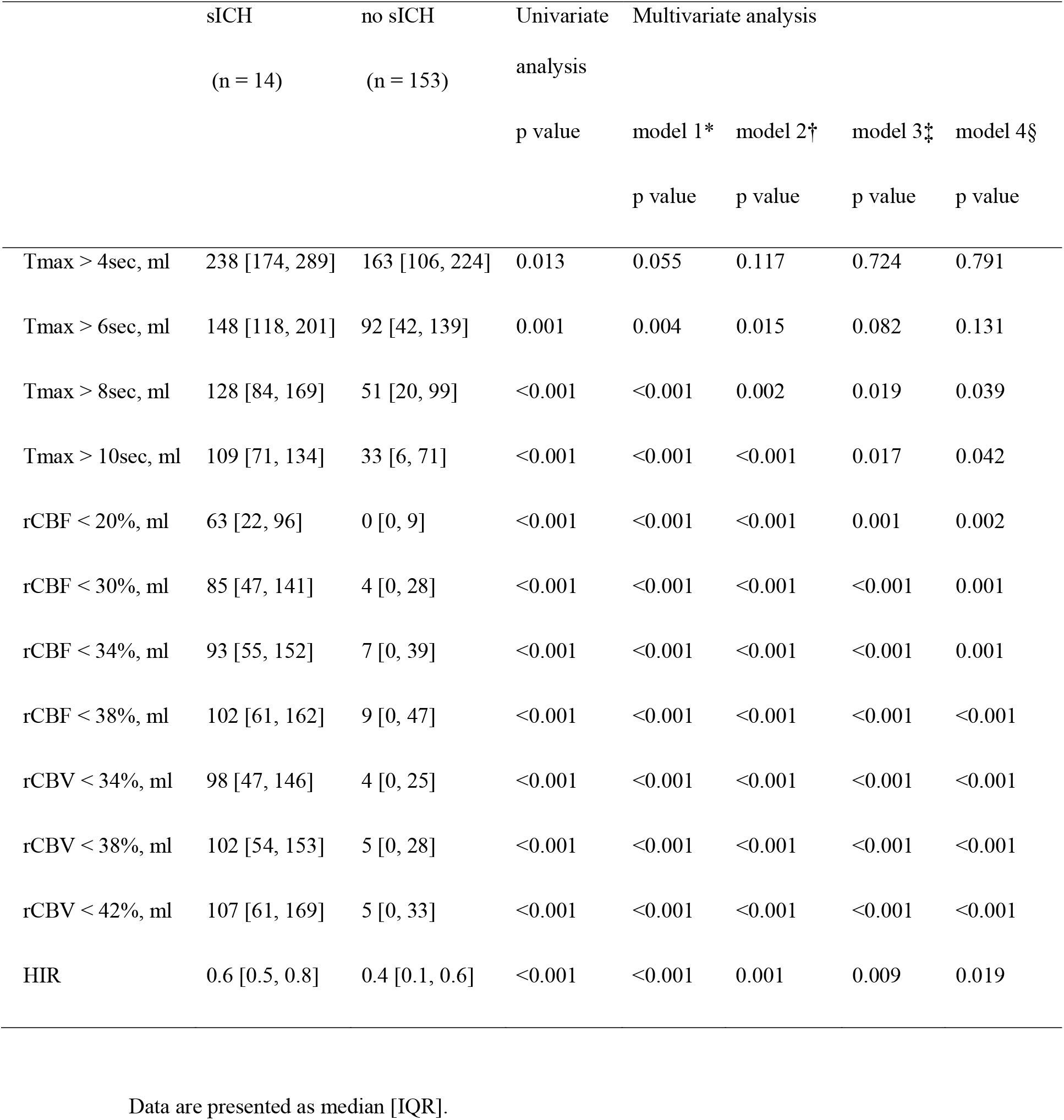
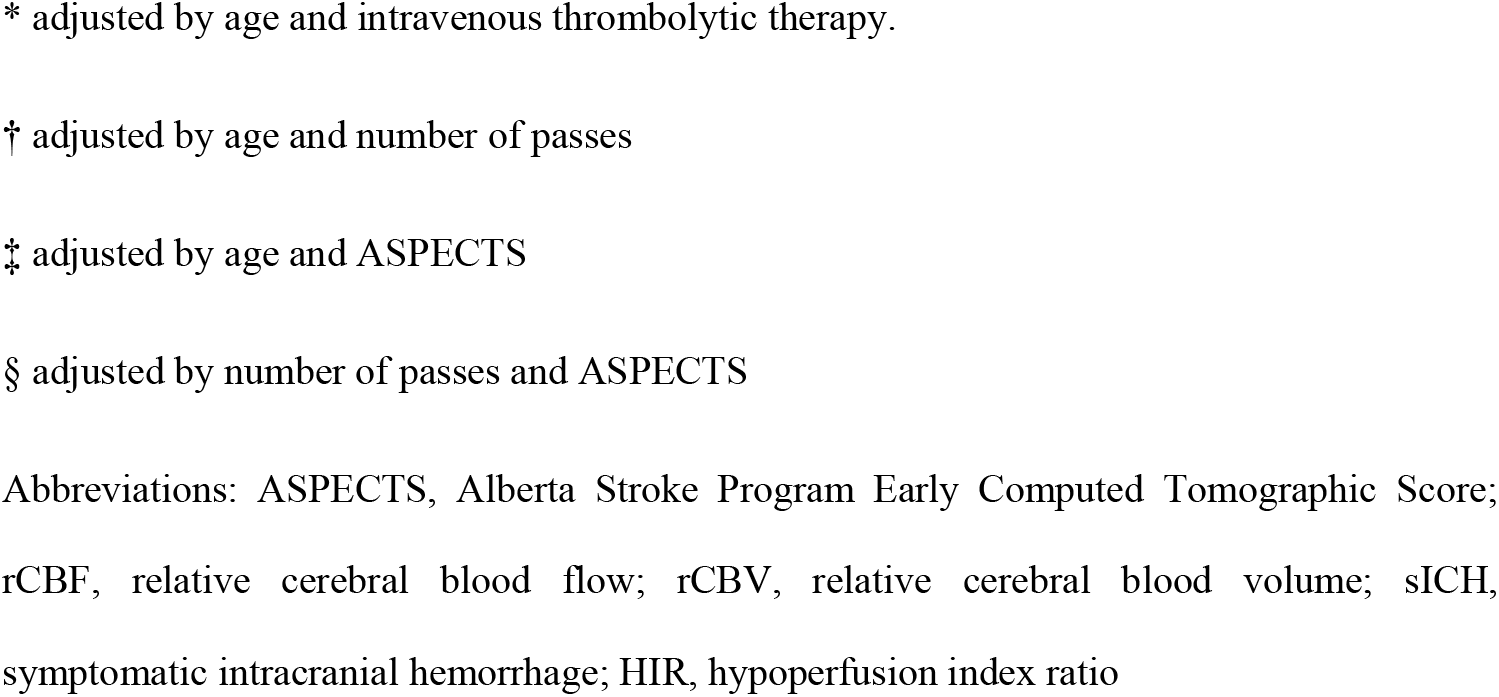
Multivariate logistic analysis of symptomatic and no symptomatic intracranial hemorrhage and CT perfusion parameters.

|  | sICH<br>(n = 14) | no sICH<br>(n = 153) | Univariate<br>analysis<br>p value | Multivariate analysis |  |  |  |
| --- | --- | --- | --- | --- | --- | --- | --- |
|  |  |  |  | model 1*<br>p value | model 2†<br>p value | model 3‡<br>p value | model 4§<br>p value |
| Tmax > 4sec, ml | 238 [174, 289] | 163 [106, 224] | 0.013 | 0.055 | 0.117 | 0.724 | 0.791 |
| Tmax > 6sec, ml | 148 [118, 201] | 92 [42, 139] | 0.001 | 0.004 | 0.015 | 0.082 | 0.131 |
| Tmax > 8sec, ml | 128 [84, 169] | 51 [20, 99] | <0.001 | <0.001 | 0.002 | 0.019 | 0.039 |
| Tmax > 10sec, ml | 109 [71, 134] | 33 [6, 71] | <0.001 | <0.001 | <0.001 | 0.017 | 0.042 |
| rCBF < 20%, ml | 63 [22, 96] | 0 [0, 9] | <0.001 | <0.001 | <0.001 | 0.001 | 0.002 |
| rCBF < 30%, ml | 85 [47, 141] | 4 [0, 28] | <0.001 | <0.001 | <0.001 | <0.001 | 0.001 |
| rCBF < 34%, ml | 93 [55, 152] | 7 [0, 39] | <0.001 | <0.001 | <0.001 | <0.001 | 0.001 |
| rCBF < 38%, ml | 102 [61, 162] | 9 [0, 47] | <0.001 | <0.001 | <0.001 | <0.001 | <0.001 |
| rCBV < 34%, ml | 98 [47, 146] | 4 [0, 25] | <0.001 | <0.001 | <0.001 | <0.001 | <0.001 |
| rCBV < 38%, ml | 102 [54, 153] | 5 [0, 28] | <0.001 | <0.001 | <0.001 | <0.001 | <0.001 |
| rCBV < 42%, ml | 107 [61, 169] | 5 [0, 33] | <0.001 | <0.001 | <0.001 | <0.001 | <0.001 |
| HIR | 0.6 [0.5, 0.8] | 0.4 [0.1, 0.6] | <0.001 | <0.001 | 0.001 | 0.009 | 0.019 |

Data are presented as median [IQR].
\* adjusted by age and intravenous thrombolytic therapy.
† adjusted by age and number of passes
‡ adjusted by age and ASPECTS
§ adjusted by number of passes and ASPECTS
Abbreviations: ASPECTS, Alberta Stroke Program Early Computed Tomographic Score; rCBF, relative cerebral blood flow; rCBV, relative cerebral blood volume; sICH, symptomatic intracranial hemorrhage; HIR, hypoperfusion index ratio

**Figure 2.**
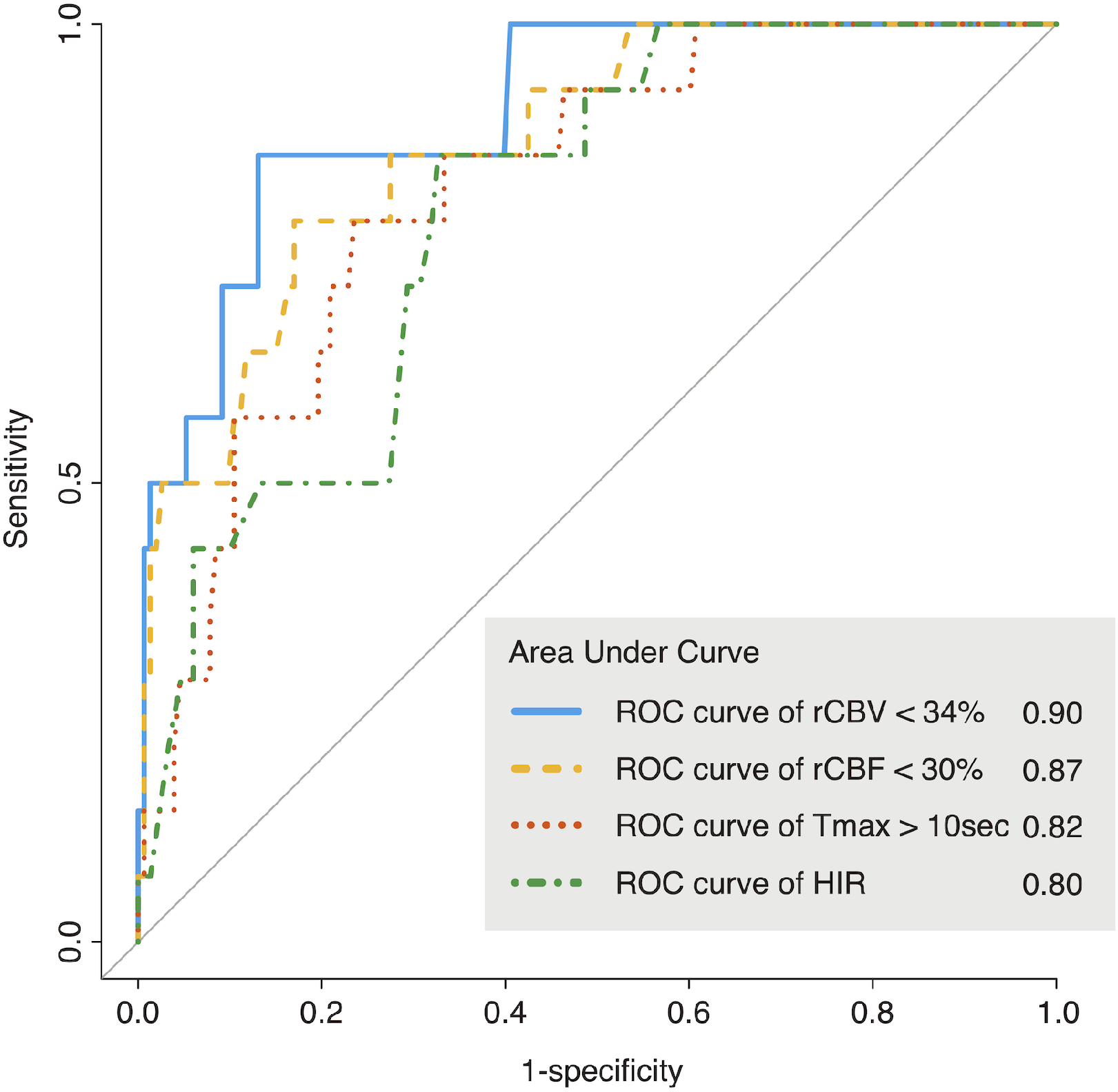
Receiver operating characteristic curve prediction ability of symptomatic intracranial hemorrhage based on rCBV < 34%, rCBF < 30%, Tmax > 10sec and hypoperfusion index ratio calculated by CT perfusion. Abbreviations: AUC, area under curve; rCBF, relative cerebral blood flow; rCBV, relative cerebral blood volume; CI, confidence intervals; sICH, symptomatic intracranial hemorrhage; HIR, hypoperfusion index ratio.

Supplemental Table 3 shows the univariate and multivariate logistic analyses between any ICH and CT perfusion parameters. After adjusting for age, intravenous thrombolytic therapy number of passes, and ASPECTS, most of parameters were correlated with any ICH other than Tmax > 4 sec and Tmax > 6 sec. The AUC of perfusion imaging for any ICH was 0.63 for Tmax > 10 sec, 0.70 for rCBF < 30%, and 0.72 for rCBV < 34%, all of which were lower than the AUC for sICH (Supplemental Table 4). Focusing on etiology, the value of Tmax > 10 sec and HIR were significantly larger in the CE cases (Supplemental Table 5). There were a few patients with sICH despite a low rCBV (Figure 3), and LAA was more common in the low rCBV group (2 (100.0%) vs 0 (0%)) (Supplemental Table 6).

**Figure 3.**
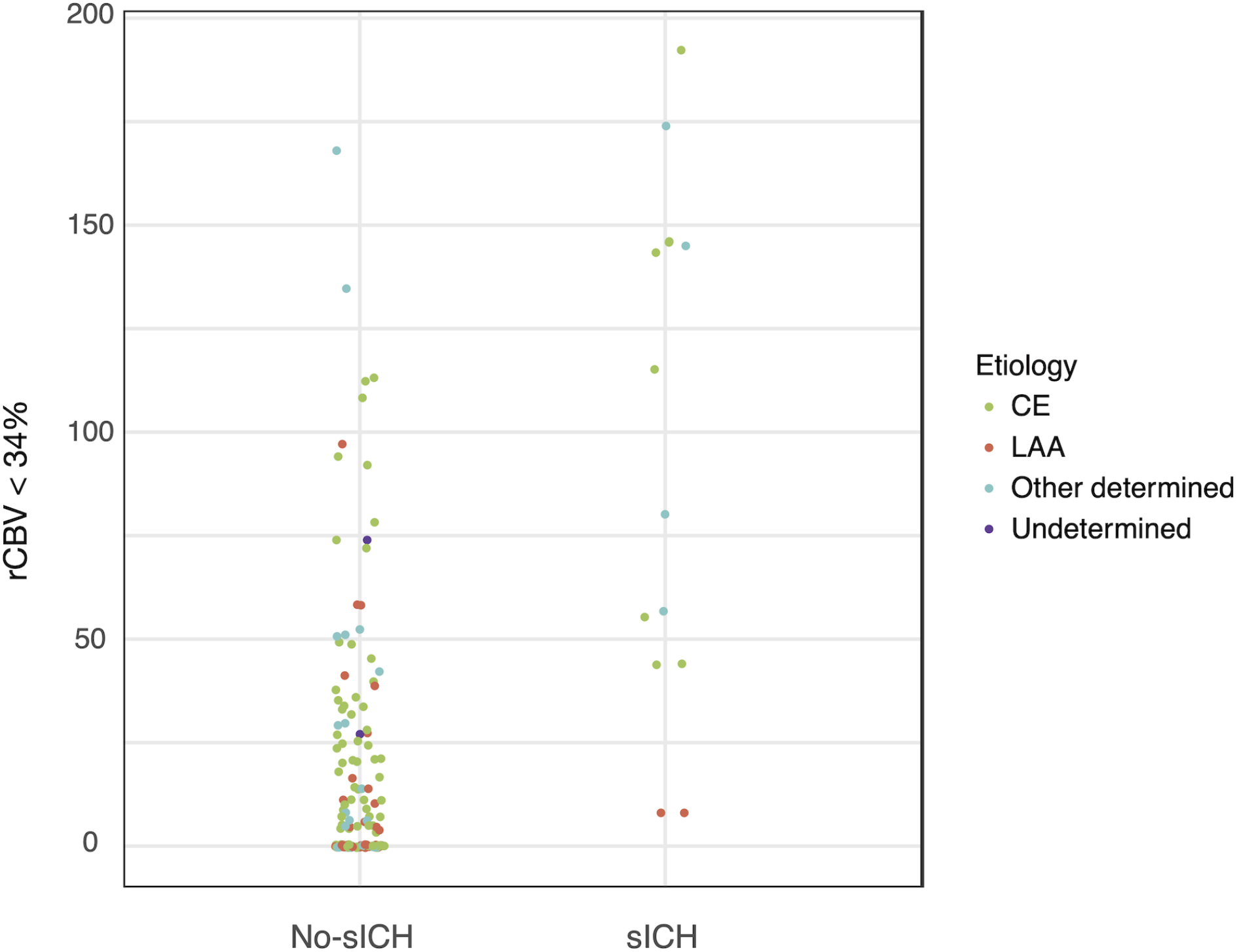
Distribution of the value of rCBV < 34% by etiology with and without symptomatic intracranial hemorrhage. Abbreviations: CE, cardiac embolism; LAA, large-artery atherosclerosis; rCBV, relative cerebral blood volume; sICH, symptomatic intracranial hemorrhage.

## Discussion

In this present study, we compared the incidence and association of HT and with the CT perfusion parameters just before MT in patients with cerebral large vessel occlusion. We have shown that the value of rCBF, rCBV, and Tmax delay were the risk factors of any ICH and sICH. Among these parameters, we have verified that rCBV is the strongest predictive factor for sICH after MT. Additionally, the patients with sICH despite the preserved rCBV were mostly diagnosed as LAA at discharge.

We compared the patients not only between the sICH and no-sICH group but also between any ICH, a-sICH and no ICH patients. Our results showed that the patients with a-sICH and sICH had significantly different background characteristics. Among the three groups, the a-sICH patients had higher pre-stroke mRS and longer procedural time. On the other hand, the sICH patients had shorter procedural time and non-significantly higher pre-stroke mRS. In other words, the patients with poor preoperative conditions were more likely to have a-sICH, and the factors were different from those causing sICH.

In previous reports, both rCBV and rCBF have been reported to be associated with HT; however, it is unclear which parameter is a stronger predictor^32,33,34^. Additionally, the association of perfusion parameters with HT after MT has not yet been well investigated, although several radiographical predictors were reported to be associated with HT in AIS^35^. Ueda et al reported the relationship between the risk of HT and reduced residual CBF after local intraarterial thrombolytic therapy^32^. However, recently, CBV was also reported to be a predictor of HT in acute ischemic stroke^36^. CBV reflects the amount of blood volume in brain tissues, and areas of decreased CBV may have greater ischemic depth than areas of decreased CBF. Although CBF is more likely to correlate with DWI lesion than CBV^22^, CBV can also reflect cerebrovascular reversible capability^37,38,39,40^; therefore, it may have a strong correlation with HT. If the parameter value of CBV is high, the hemorrhagic complication may be indicated after successful recanalization. Yu et al. reported that the regional pial collateral flow predicts the occurrence of HT and correlates most closely with rCBV among the CT perfusion parameters^41^. Nevertheless, in the present study, rCBF is also a strong predictor of sICH, which is consistent with a previous report showing that the patients with a large ischemic core are at a high risk of HT. Patients with an infarct core of > 53 ml in CBF <30% lesion reportedly had poor outcomes, even within 3 hours from disease onset, they are defined as a malignant profile^42^. As in previous reports, both CBF and CBV are strong predictors of sICH; however, rCBV was found to be a stronger predictor in this study.

In our cohort, there were two patients of sICH despite a preserved CBV; both patients had LAA and the ratio of LAA was higher than that of the other group. We speculated three reasons for these pathophysiological differences. In case of LAA, chronic low perfusion is present^43,44^; therefore, a hyper-perfusion phenomenon after MT occurs more frequently than CE. Second, dual antiplatelet therapy after MT is frequently performed in LAA patients, which may cause hemorrhagic changes. Third, blood pressure may be more strictly controlled when the stenotic lesion is observed after MT because such a cases are at risk for re-occlusion. However, due to the small number of patients, it is difficult to suggest an association between LAA and ICH. This point needs to be investigated in future studies with a larger number of patients.

There are several limitations in our study. First, this present study was a single-center and retrospective cohort study. However, in the patients with malignant profiles as large cores, recanalization is not recommended because of the risk for HT. Furthermore, only a few facilities have an automated perfusion analysis system in Japan. Therefore, a perfusion-related RCT is difficult to actually conduct. Second, the number of patients with adequate perfusion imaging was small in this population. CT perfusion was not performed in 31 patients mainly due to renal dysfunction, and it was not calculated accurately in 24 patients due to a low cardiac function, the problem of venous contrast route, and motion artifact. This can be a limitation of perfusion imaging itself. It may be necessary to change the setting of CT perfusion, such as increasing the post contrast time to reduce the error during future RCT. Finally, there were some cases of large ischemic core detected by non-contrasted CT, and we did not perform any reperfusion therapy to these cases; these cases were not enrolled in our cohort, and it is unclear whether reperfusion therapy could be harmful owing to hemorrhagic change in these patients.

In conclusion, the high value of rCBV < 34% was the most predictive parameter for sICH after MT. Stroke subtypes such as LAA was more frequently occurring sICH even not having an imaging malignant profile.

## Data Availability

The data supporting this study's findings can be accessed from the corresponding author upon reasonable request.

## Non-standard abbreviations and acronyms

ICH: intracranial hemorrhage
AUC: area under the curve
a-sICH: asymptomatic intracranial hemorrhage
CE: cardiac embolism
HT: hemorrhagic transformation
HIR: hypoperfusion intensity ratio
IQR: interquartile range
LAA: large-artery atherosclerosis
NIHSS: National Institutes of Health Stroke Scale
MT: mechanical thrombectomy
mRS: modified Rankin Scale
mTICI: modified Thrombolysis in Cerebral Infarction
RCTs: randomized controlled trials
ROC: receiver operating characteristics
rCBF: relative cerebral blood flow
rCBV: relative cerebral blood volume
Tmax: time of maximum concentration
TOAST: Trial of Org 10172 in Acute Stroke Treatment
sICH: symptomatic intracranial hemorrhage

## Acknowledgements

Dr. Daisuke Sakamoto, Dr. Shun Ono, Dr. Shuntaro Kuwahara, Dr. Taiki Chida, Dr. So Sawamura (Hyogo Medical University), and Dr. Yoichiro Nagao (Saiseikai Kumamoto Hospital) helped with data collection. Ms. Yuko Nagai, Ms. Yui Kurosaka, and Ms. Iori Matsunaga contributed to the secretarial work.

## Supplemental Material

Supplemental Tables 1–6

STROBE checklist

---

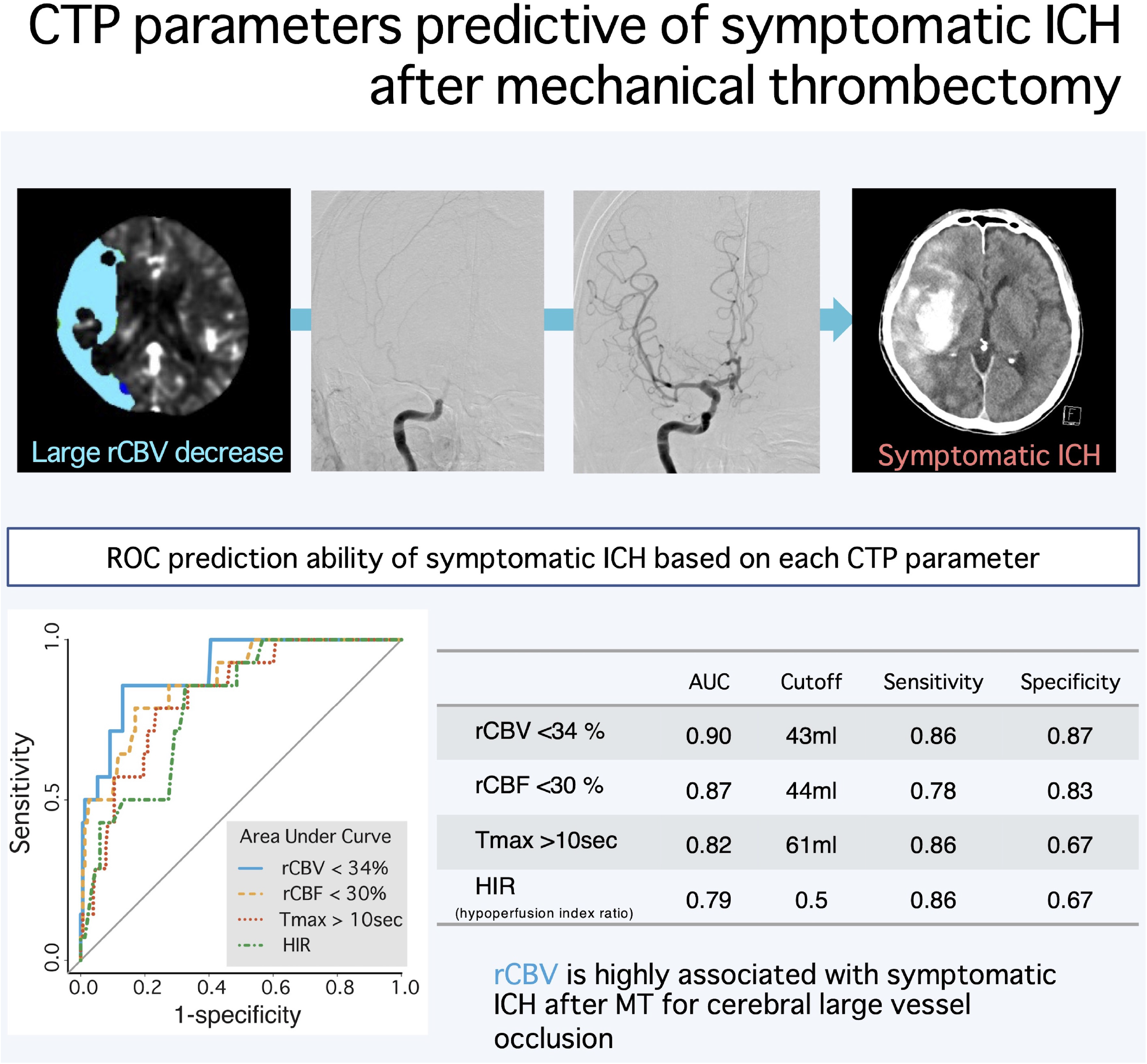

